# Methylome and transcriptome functional analysis identifies key biomarkers in ketamine’s sustained therapeutic effect on PTSD

**DOI:** 10.1101/2025.05.25.25327632

**Authors:** Nathan J. Wellington, Ana P. Bouças, Paul E. Schwenn, Jim Lagopoulos, Bonnie L. Quigley, Anna V. Kuballa

## Abstract

**Background:** Ketamine has emerged as a rapid-acting therapeutic option for post-traumatic stress disorder (PTSD); however, its ability to sustain long-term therapeutic outcomes remains poorly understood. Identifying molecular biomarkers predictive of a sustained response to ketamine may enhance personalised treatment strategies. This study investigates the epigenetic and transcriptomic precursors underpinning ketamine’s long-term therapeutic efficacy in PTSD.

**Methods:** This study utilised data from the Oral Ketamine Trial of PTSD (OKTOP), an open-label, dose-ranging clinical study conducted between 2021 and 2024. Baseline differential DNA methylation and gene expression profiles of sustained responders (clinical response >1 month post-ketamine treatment) were compared against non-responders. Epigenomic and transcriptomic analyses were performed to identify differentially regulated genes associated with ketamine response.

**Results:** Baseline molecular analyses revealed significant differential methylation and transcriptomic profiles across 112 genes. Key biomarkers included DENND5B (cg02046589), ZFY (cg00272582), PDGFRA (cg21309167), CPT1A (cg10098373), AHRR (cg26076054), RPH3AL (cg17316718), CHI3L1 (cg19081101), UTY (cg04790916), LDHD (cg00004883), TBC1D16 (cg26287152), FAM66A (cg23285059), NME8 (cg02531859), EIF1AY (cg13308744), PCBP3 (cg13695288), PAQR6 (cg03954786), KCNK17 (cg19475903), PLPP2 (cg24452451), ANK1 (cg23668222), LINC00200 (C10ORF139, cg19282259), ALAS2 (cg07471703), ZBP1 (cg06305758), TACSTD2 (cg01821018), and PLEKHH3 (cg24455236). These biomarkers were implicated in pathways related to metabolism, transcriptional regulation, cell signalling, neuronal development, immune response, synaptic plasticity, and cytoskeletal organisation. Non-responders exhibited persistent dysregulation across these pathways, suggesting potential biological barriers to treatment efficacy. Clinically, sustained responders presented with higher baseline PTSD severity and demonstrated a response at lower ketamine doses compared to non-responders.

**Conclusions:** This study highlights the potential of methylomic and transcriptomic profiling to identify functional biomarkers predictive of ketamine response in PTSD. The observed molecular distinctions between responders and non-responders suggest a complex interplay between clinical presentation and treatment outcomes. These findings contribute to advancing precision medicine approaches for PTSD by informing biomarker-driven treatment stratification and optimisation of ketamine therapy.

## 1. Introduction

Post-traumatic stress disorder (PTSD) is a debilitating mental health condition characterised by intrusive memories, heightened arousal, and emotional numbness following exposure to traumatic events. Traditional pharmacological interventions for PTSD, such as selective serotonin reuptake inhibitors (SSRIs) and cognitive-behavioural therapy (CBT), have demonstrated limited efficacy, especially in individuals with chronic or treatment-resistant PTSD, with findings revealing a low response rate of 30-50% (1, 2). This has led researchers to explore alternative treatments, with ketamine emerging as a promising candidate due to its rapid onset of action and potential to mitigate the symptoms of PTSD for sustained periods (3).

Ketamine, originally used as an anaesthetic, has garnered attention for its off-label use as a rapid-acting antidepressant and promising treatment for PTSD (4). Unlike conventional antidepressants that can take weeks to show effects, ketamine in sub-anaesthetic doses has been shown to produce rapid improvements in mood and symptoms of PTSD within hours of administration(5). While the acute effects of ketamine on PTSD symptoms are well-documented, the mechanisms underlying its sustained efficacy over weeks and months remain poorly understood(6, 7). A growing body of research suggests that the lasting effects of ketamine may be mediated, in part, through epigenetic induced changes in gene expression(8, 9). Understanding these epigenetic and gene expression changes will be vital to maximising ketamine’s therapeutic potential in PTSD.

Epigenetic changes, specifically DNA methylation, play a pivotal role in regulating gene expression, particularly in response to environmental stressors such as trauma(10). PTSD has been shown to induce widespread changes in methylation patterns, which can have long-term effects on the regulation of genes involved in stress response, immune function, and neural plasticity(11). Further, studies have found that long-term methylation changes in key neurotransmitters in adulthood have been associated with childhood adversity(12, 13). Yet little is understood as to how these altered methylation patterns influence treatment response.

Recent epigenome-wide studies have repeatedly identified differential methylation positions (DMP) specifically linked to PTSD susceptibility in important gene sites including IGHJ3P, PDE9A, MLH1, AHRR, G0S2, IL-6, USP49, C19of55, AIM2, GRIN2A, C21of56, SKA2, MET and MAD1l1(11, 14–22). These sites are yet to be explored with regard to ketamine’s influence on their methylation patterns and gene expression, providing an opportunity to identify whether methylation response patterns renormalise following ketamine therapy.

This study builds on the findings of the Oral Ketamine Trial of PTSD (OKTOP), an open-label clinical trial conducted between July 2021 and January 2024. The OKTOP study aimed to explore the efficacy, feasibility, and tolerability of oral ketamine in treating PTSD(23). Participants were administered escalating doses of oral ketamine over six weeks, with follow-up assessments conducted at 10 weeks to evaluate the durability of treatment effects. The trial demonstrated that while some participants showed a sustained response to ketamine, others did not, highlighting the heterogeneity in treatment outcomes. The aim of this investigation was to explore the underlying molecular mechanisms that differentiate ketamine sustained responders from non-responders, with a specific focus on epigenetic and gene expression biomarkers before treatment.

This study holds the potential to contribute significantly to the growing field of biomarker discovery in PTSD. Biomarkers, particularly those related to gene expression and epigenetic modifications, offer a promising avenue for the development of personalised treatment strategies. In the context of PTSD, where treatment resistance is common and therapeutic options are limited, the identification of biomarkers that can predict treatment response is of paramount importance. The integration of epigenetic and gene expression data will provide a more nuanced understanding of the biological precursors and their functional relevance that may contribute to how ketamine induces long-lasting changes in neural circuits and behaviour.

This knowledge is crucial for developing novel therapeutic strategies that can mimic the beneficial effects of ketamine while minimising its potential side effects.

## 1. Methods

### 1.1 Study Design

This study builds on the OKTOP study conducted between July 2021 and January 2024, approved by the Prince Charles Hospital (PCH) Human Research Committee (HREC/18/QPCH/288) and University of the Sunshine Coast (UniSC) Ethics Committee (A181190) (23). Genetic analysis and reporting were part of the Genetic Biomarkers of Ketamine on PTSD (GBOK) project, with ethics approvals from PCH (HREC/18/QPCH/288) and UniSC (S211655). The OKTOP study was an open-label, dose-ranging clinical trial that assessed the efficacy, feasibility, and tolerability of oral ketamine for PTSD treatment. Participants underwent a 10-week trial, including six weeks of active treatment and two follow-up assessments. Clinical evaluations and blood samples were collected at baseline (BAS), mid-treatment (Week 3), final treatment (Week 6), one-week post-treatment (Week 7, FUP1), and four weeks post-treatment (Week 10, FUP2). The study recruited adult participants (aged 18 and above) diagnosed with PTSD, confirmed through the Clinician-Administered PTSD Scale for DSM-5 (CAPS-5)(24). Baseline assessments, including physical examination, urinalysis, clinical blood tests, and medical history, were completed within 14 days before starting ketamine treatment.

Participants received a sub-anaesthetic dose of oral ketamine weekly, monitored by a psychiatrist and health clinician. The initial dose was set at 0.5 mg/kg and gradually increased by 0.1-0.5 mg/kg each week, based on individual tolerance, to a maximum dose of 3.0 mg/kg.

### 1.2 Participants

Participants included within this study (*n* = 19, 7M/ 12F) were Caucasian, with an age range of 43 to 77 years (*M* = 57.8yrs, *SD* = 7.6) (**Table 1**). The study participants were divided into outcome groups based on the study definition of treatment response: non-responders were participants that did not achieve and maintain a ≥50% reduction in PTSD Checklist for DMS-5 (PCL-5) score from BAS to FUP1 and FUP2 assessments (*n* = 8) while sustained responders achieved and maintained a ≥50% PCL-5 score reduction at both FUP timepoints (*n* = 11). The mean age was similar between the outcome groups, with non-responders (*M* = 48.68 years, *SD* = 11.23) and sustained responders (*M* = 50.69 years, *SD* = 14.21), *t*(17) = -0.34, *p* = 0.665, indicating no significant differences in age. Likewise, the average weight showed no significant variation between non-responders (*M* = 87.69 kg, *SD* = 12.98) and sustained responders (*M* = 87.05 kg, *SD* = 24.49), *t*(17) = 0.07, *p* = 0.9925.

**Table 1:** Demographic and clinical characteristics.

| Characteristic | Non-responders<br>(Mean ± SD) | Sustained<br>Responders<br>(Mean ± SD) | t-statistic | p-value |
| --- | --- | --- | --- | --- |
| **Participants** | 8 | 11 |  |  |
| **Age (years)** | 48.68 ± 11.23 | 50.69 ± 14.21 | -0.34 | 0.665 |
| **Weight (kg)** | 87.69 ± 12.98 | 87.05 ± 24.49 | 0.07 | 0.992 |
| **Sex - Female** | 5 | 7 |  |  |
| **- Male** | 3 | 4 |  |  |
| **PCL-5 BAS** | 34.13 ± 4.21 | 41.27 ± 3.75 | -3.82 | **0.001** |
| **PCL-5 FUP2** | 34.43 ± 17.99 | 13.10 ± 8.47* | 3.11 | **0.006** |
| **MADRS BAS** | 33.88 ± 3.42 | 31.64 ± 1.83 | 1.69 | 0.11 |
| **MADRS FUP2** | 12.38 ± 11.08* | 6.82 ± 5.99* | 1.29 | 0.215 |
| **DASS BAS** | 48.25 ± 8.19 | 51.09 ± 5.16 | -0.86 | 0.400 |
| **DASS FUP2** | 52.86 ± 23.58 | 19.10 ± 5.22 | 3.98 | **0.001** |
| **WHO-5 BAS** | 35.00 ± 6.92 | 24.73 ± 6.78 | 3.22 | **0.005** |
| **WHO-5 FUP2** | 37.60 ± 27.51 | 60.00 ± 24.33* | -1.84 | 0.083 |
| **Sig. Trauma Type** | Childhood (75%) | Childhood (36%) |  | 0.17<sup>†</sup> |
| **Sig. Comorbid**<br>**Diagnosis** | MDD (100%) | MDD (63%) |  | **0.02<sup>†</sup>** |

Note: \* = Non-responders /Sustained responders significant difference between timepoint clinical
results. Degrees of freedom were calculated on equal variance (all were $df(17)$ ), p-values were two-

### 1.3 Clinical Measures

The primary outcome measure was the PCL-5 from BAS to FUP2. Secondary measures included the Montgomery–Åsberg Depression Rating Scale (MADRS), Depression, Anxiety and Stress Scale (DASS-21), Beck’s Scale for Suicide Ideation (BSS), Social and Occupational Assessment Scale (SOFAS), Snaith-Hamilton Pleasure Scale (SHAPS-C), and the World Health Organisation Well-Being Index Five (WHO-5). These assessments were conducted at BAS, during the intervention, and at FUP1 and FUP2.

### 1.4 Blood Collection

Approximately 50-70 mL of whole blood was drawn by a phlebotomist. Samples were processed into whole blood, serum, plasma, and peripheral blood mononuclear cells (PBMCs). PBMCs were further separated into aliquots and stored either as a dry cell pellet, in foetal bovine serum with 10% dimethyl sulfoxide (DMSO), or with 600 μL RNeasy lysis buffer (Qiagen) and 6 μL of beta-mercaptoethanol to preserve RNA integrity. All samples were stored at -80°C.

### 1.5 DNA Methylation and RNA Sequencing Acquisition

BAS samples were analysed to assess both methylation and gene expression differences in response to ketamine exposure. A total of 19 PBMC samples suspended in lysis buffer (sustained responders *n* = 11, non-responders *n* = 8) were transported on dry ice to the Australian Genome Research Facility (AGRF) for DNA methylation and RNA sequencing analysis. DNA was extracted using the DNeasy Blood and Tissue kit (Qiagen), RNA was extracted via the RNeasy mini kit (Qiagen) as per manufacturer’s instructions for all samples.

Bisulphite conversion was conducted on the DNA samples using the Zymo EZ-96 DNA Methylation kit. Samples were analysed on an Illumina Human Methylation EPIC v2.0 Bead Chip utilising the Illumina Infinium HD Methylation assay protocol. Genome Studio v2011.1 with the Methylation module 1.9.0 software and Illumina EPIC-8v2-0_A1 manifest file was used to generate intensity data (IDAT) files. All methylation samples met the requirement of ≥96% detected CpG sites (*p*<0.05), based on non-cancer samples. Normalisation and differential analysis were performed in Genome Studio v2011.1, adjusting for false discovery rate (FDR) using Illumina’s differential error model and a difference score applied. The data has been deposited in the GEO database under accession number GSE287261.

Gene expression analysis was conducted using a NovaSeq utilising the Illumina RNA-Seq 20 million paired end sequencing protocol. Image analysis was carried out in real-time using NovaSeq Control Software (NCS) v1.2.0.28691 and Real-Time Analysis (RTA) v4.6.7 on the instrument computer. The Illumina DRAGEN BCL Convert pipeline (07.021.645.4.0.3) was used to produce sequence data. The data underwent quality control, trimming, alignment, annotation, and differential expression analysis using CLC Genomics Workbench 25.0 (Qiagen). Reads were mapped to the homo sapiens genome assembly GRCh38.p14, RefSeq release 227 and CCDS release 24 databases for annotation. The data has also been published in the GEO database under accession number GSE288174.

### 1.6 Statistical Analysis

All BAS samples were analysed utilising R Studio 2024.04.2 Build 764 incorporating limma, ggplot2, dply and minifi packages(25–29). P-values were calculated on the methylation data through the Wilcoxon signed-rank test for each CpG site. These values were then FDR-adjusted using the Benjamini-Hochberg method and p-values were log10 transformed. Fold changes were calculated from the difference between average beta scores and log-2 calculations were applied. The summation of the ranked data was determined according to their p-value, fold change and difference score.

### 1.7 Pathway Analysis

Pathway analysis was conducted utilising Ingenuity Pathway Analysis 24.0.1 (Qiagen). Methylation data was imported applying p-value and fold change parameters. CpG islands were mapped through the Illumina Human Methylation, University of California Santa Cruz Genomics Institute Jan 2022 (USCS) and GENECODE release 47 databases to filter CpG sites related by position (island, shore or shelf) as to their influence on the gene, with CpG islands outside these positions excluded. RNA seq data was imported and mapped to HUGO Gene Nomenclature Committee (HGNC) and RefSeq release 227 databases, applying their p-value and expression fold change. Both databases were analysed through the core analysis function applying a p-value of <0.05 and RNA-Seq data was interpreted through fold change ≤ -1 or ≥ 1.

## 2 Results

### 2.1 Clinical Characteristics

BAS and FUP2 clinical assessments highlighted several notable distinctions between groups. The average baseline PCL-5 score, a measure of PTSD symptom severity, was significantly higher in the sustained responders to ketamine group (*M* = 41.27, *SD* = 3.75) compared to non-responders (*M* = 34.13, *SD* = 4.21), *t*(17) = -3.82, *p* = 0.001. Additionally, the MADRS baseline scores, reflecting depression severity, were slightly higher in the non-responder cohort (*M* = 33.88, *SD* = 3.42) compared to the sustained responder cohort although not significant (*M* = 31.64, *SD* = 1.83), *t*(17) = 1.69, *p* = 0.11. The WHO-5 score, reflecting psychological well-being, was significantly lower among sustained responders (*M* = 24.73, *SD* = 6.78) compared to non-responders (*M* = 35.00, *SD* = 6.92), *t*(17) = 3.22, *p* = 0.005, suggesting differential baseline well-being levels. FUP2 clinical assessments, performed four weeks post treatment completion, demonstrated significant differences between non-responders and sustained responders PCL-5 scores, which on average dropped by 28 points (*M* = 13.10, *SD* = 8.47), noting clinically meaningful improvements in symptoms while the non-responders remained within the equivalent threshold of their BAS assessments. Further clinical improvements included the MADRS scores, whereas both cohorts reported significant reductions in symptom severity, non-responders showed a reduction by 21 points (*M* = 12.38, *SD* = 11.08) and sustained responders exhibited a reduction by 25 points (*M* = 6.82, *SD* = 5.99), thus qualifying this cohort as no longer experiencing depressive symptoms.

The prevalence of significant childhood trauma differed markedly between groups, with 75% of non-responders and 36% of responders reporting childhood trauma as the primary traumatic experience. Furthermore, all non-responders were diagnosed with major depressive disorder (MDD), whereas this diagnosis was present in 63% of the sustained responders.

This study also found that participants who responded well to ketamine generally received a lower average dose compared to non-responders. Sustained responders (*M* = 1.78, *SD* = 1.2) had a final titration dose that was 31.54% lower than that of non-responders (*M* = 2.6, *SD* = 0.64), *t*(17) = 1.92, *p* = 0.07. (**Fig.1, Supplemental Material Table 1**).

**Fig.1.**
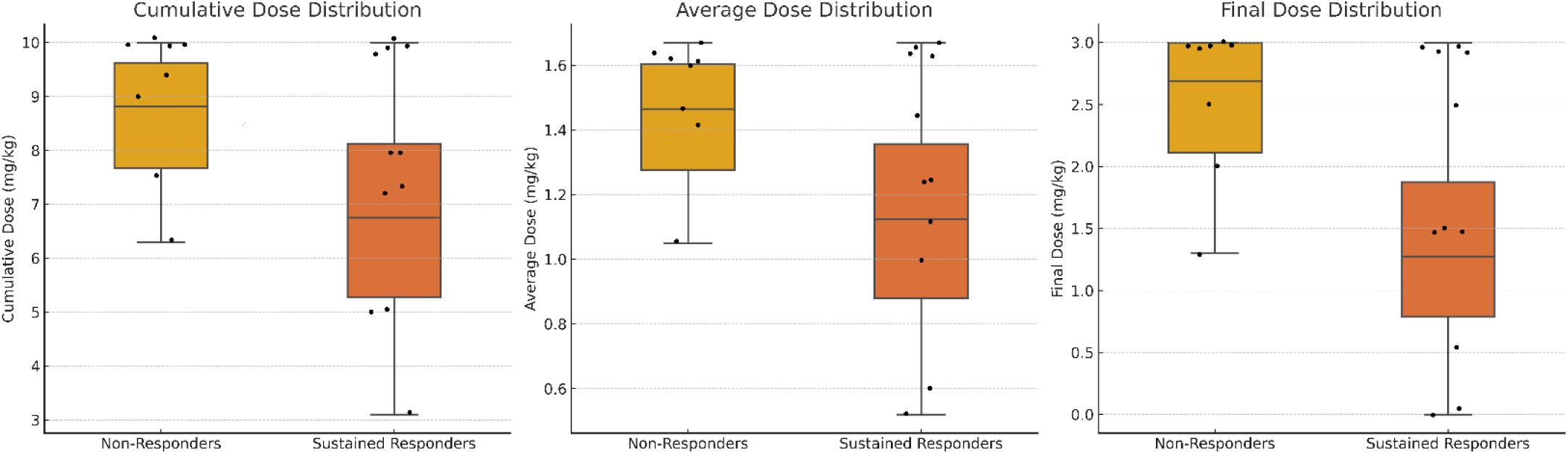
Ketamine mg/kg dosage comparisons between sustained responders and non-responders. These box and whisker plots compare non-responders (orange) and sustained responders (red) across three dose metrics: Cumulative Dose, Average Dose, and Final Dose, all measured in mg/kg. Each plot illustrates the median, interquartile range, data range excluding outliers, and individual data points. Cumulative Dose represents the total administered dose, Average Dose shows the mean dose per individual, and Final Dose indicates the last administered dose.

### 2.2 Methylome and Transcriptome Analysis

To identify key biomarkers, differential methylation analysis compared non-responders to sustained responders at BAS across 936 991 potential CpG islands, including single nucleotide polymorphisms (SNPs) that may affect methylation levels due to allelic differences. Data analysis using a p-value of ≤ 0.05 and a difference score of >13 for significance identified 27 417 CpG islands that were considered significantly differentially methylated between the response groups. Interestingly, none of the SNPs investigated were found to be significant. Of the significant CpG islands, 777 mapped to an island, shore or shelf region either upstream or downstream of genes within the GRCh38.p14 genome.

Differential gene expression analysis was performed comparing non-responders to sustained responders from a database of 59 116 potential RNA expression points, including long noncoding RNAs, complementary RNAs and mitochondrial RNAs. Data analysis using a p-value of ≤ 0.05 and expression fold changes <-1.0 and >1.0 identified 3 918 genes that were significantly differentially expressed, with 1 565 higher and 2 353 lower in sustained responders, genes.

On comparing the intersection between significant results from the methylation and gene expression data, an overlap of 112 genes was identified (**Fig.2, Supplemental Material Table 2**).

**Fig.2.**
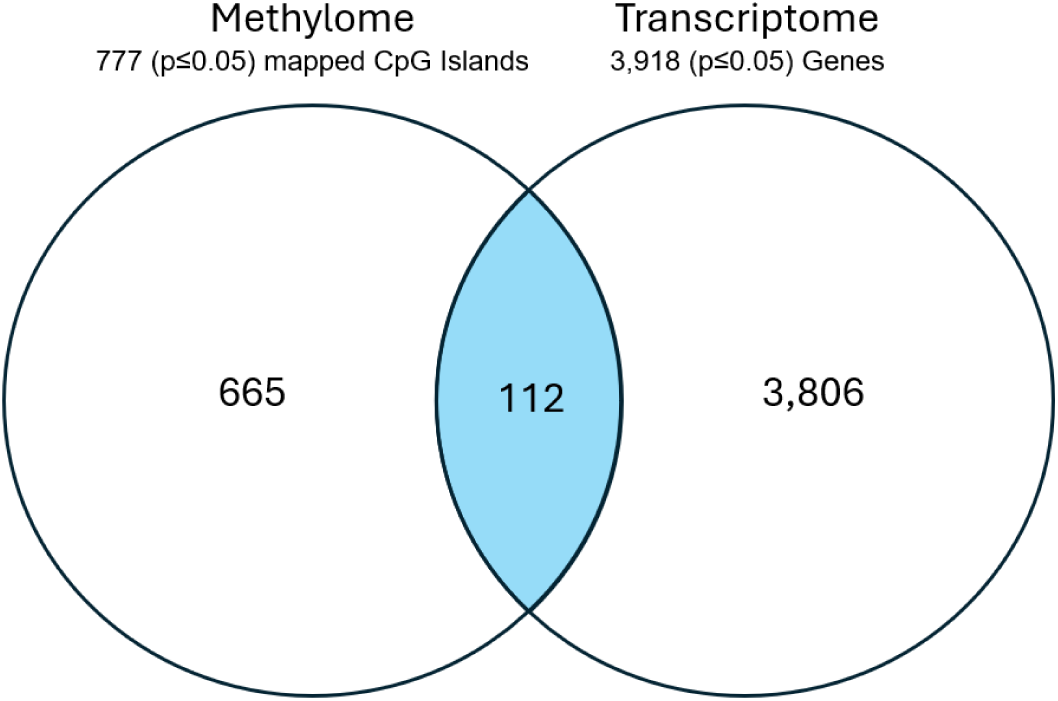
Venn diagram of significant differential baseline changes between the methylome and transcriptome of non-responders compared to sustained responders. Genes identified to have significantly higher/lower CpG island methylation in island, shore or shelf regions of the gene between sustained and non-responders at baseline are represented by the methylome results. Genes identified to have significant differential gene expression (higher or lower) between sustained and non-responders at baseline are represented by the transcriptome results.

These results highlight the influence of methylation patterns on gene expression when comparing non- and sustained responders.

Distinct patterns of methylation were identified in the 112 gene set (**Supplemental Material Table 2**) common to both methylome and gene expression analysis, with 31 genes that were hypermethylated and associated with a reduced level of gene expression (e.g. DENN5B (*fc* = -1.18) cg02046589 (Δbeta = 0.146) and ZFY (*fc* = -1.407) cg00272582 (Δbeta= 0.15)) while 42 were hypomethylated and associated with increased gene expression (e.g. CHI3L1 (*fc* = 1.521) cg19081101 (Δbeta = -0.146)). The remaining genes (*n* = 39) deviated from the typical inverse relationship between gene expression and DNA methylation (e.g., PDGFRA (fc = - 1.405), cg21309167 (Δbeta = -0.203)). This observation implies that clustering methylome and transcriptome data into cohorts may yield results that, despite being significant for individual molecules, do not conform to the established methylation-gene expression paradigm, where increased methylation is generally associated with decreased gene expression. This is an important distinction when evaluating biomarker potential. Figures 3a and b illustrate the methylation patterns of hypo- and hyper-methylation and their corresponding expression influences (**Fig. 3a & b, Supplemental Material Table 2**).

**Fig.3.**
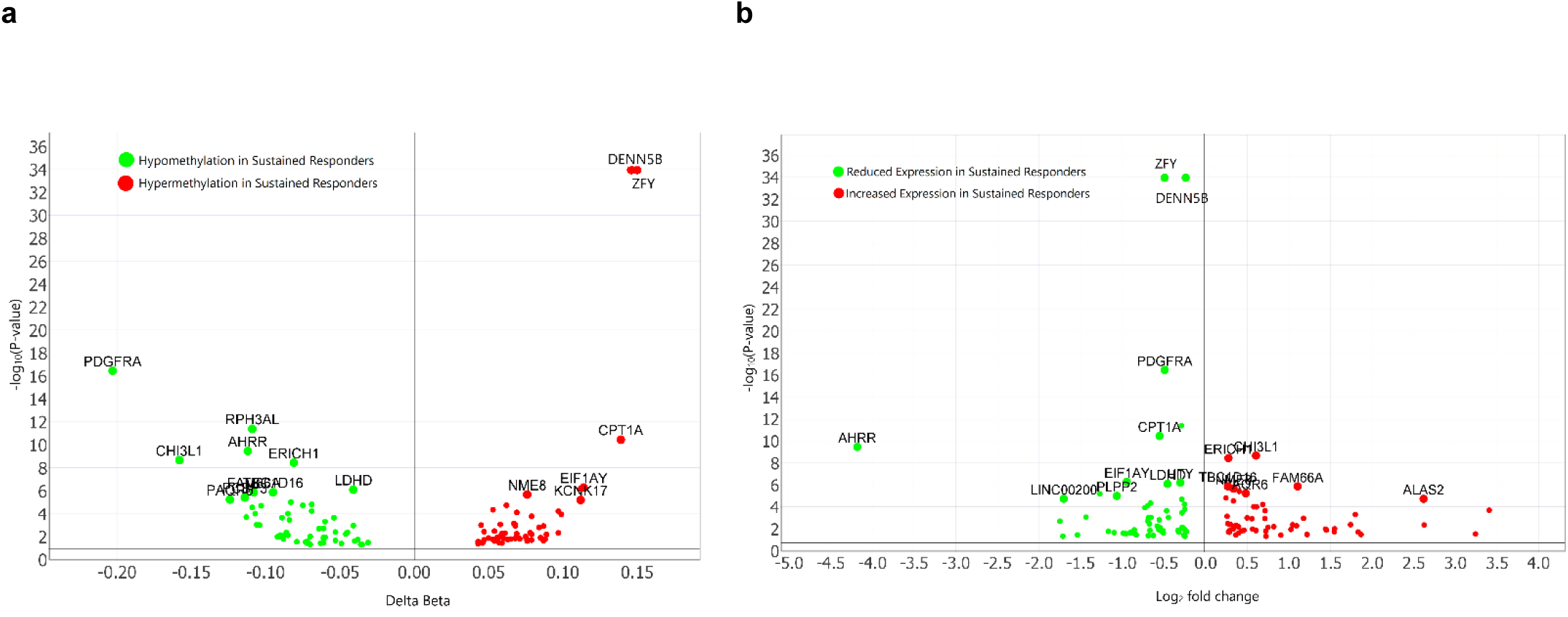
Differential DNA methylation (a) and gene expression plots (b) comparing sustained and non-responders to ketamine. These volcano plots illustrate the differential methylation and transcriptome profiles, respectively, between sustained and non-responders prior to ketamine treatment. **Fig.3a** (DNA Methylation Volcano Plot) displays the log₂ fold change (Δβ) on the x-axis, representing the direction and magnitude of methylation differences, while the y-axis shows the -log₁₀(p-value), indicating statistical significance. Genes with significant hypomethylation are highlighted in green, and genes with significant hypermethylation are indicated in red. The annotated genes represent genes with a corresponding CpG influence over p≤ 5 × 10-5. **Fig.3b** (Gene Expression Volcano Plot) highlights changes in RNA expression, with the x-axis representing the expression log ratio and the y-axis displaying the -log₁₀(p-value). The annotated genes represent differential gene expression over p≤ 5 × 10^-5^. The corresponding genes with significantly reduced gene expression are shown in green, while those with significantly increased gene expression are shown in red. By combining the methylation and transcriptome data, patterns of influence in notable genes, including DENND5B cg02046589, ZFY cg00272582, CPT1A cg10098373, and PDGFRA cg21309167 (labelled) reflect their potential as predictive biomarkers in their differential profile which may influence the participants response to ketamine treatment. Together, these plots underscore the interplay between methylation and transcriptional differences of genes with a potential role in influencing treatment outcomes.

Genes with CpG significance greater than (p≤ 5 × 10^-5^) are summarised in table 3. The table includes the methylation beta, and respective gene expression fold change values alongside their corresponding molecular function mapped using the Gene Ontology database(30).

**Table 3:**
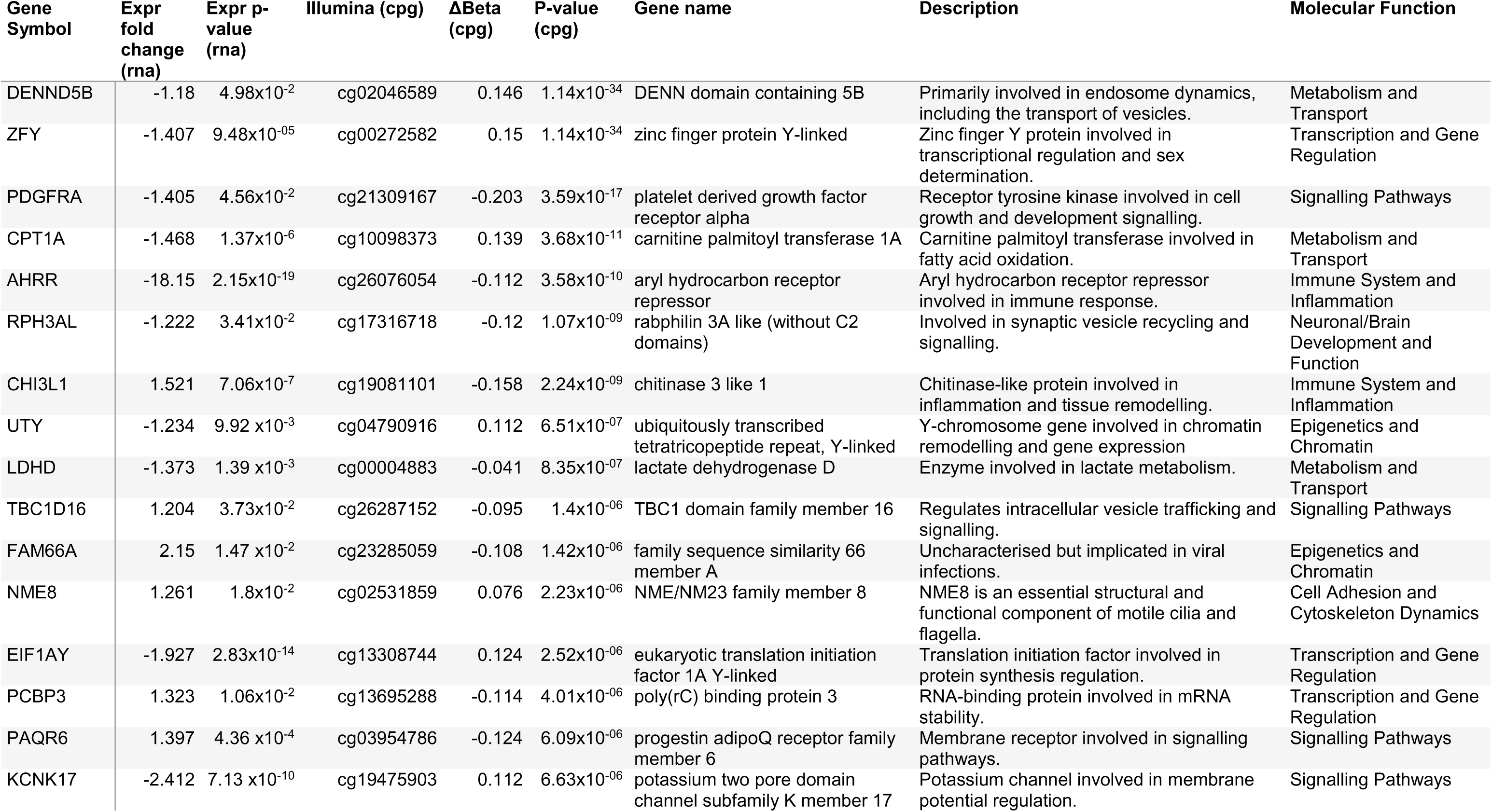
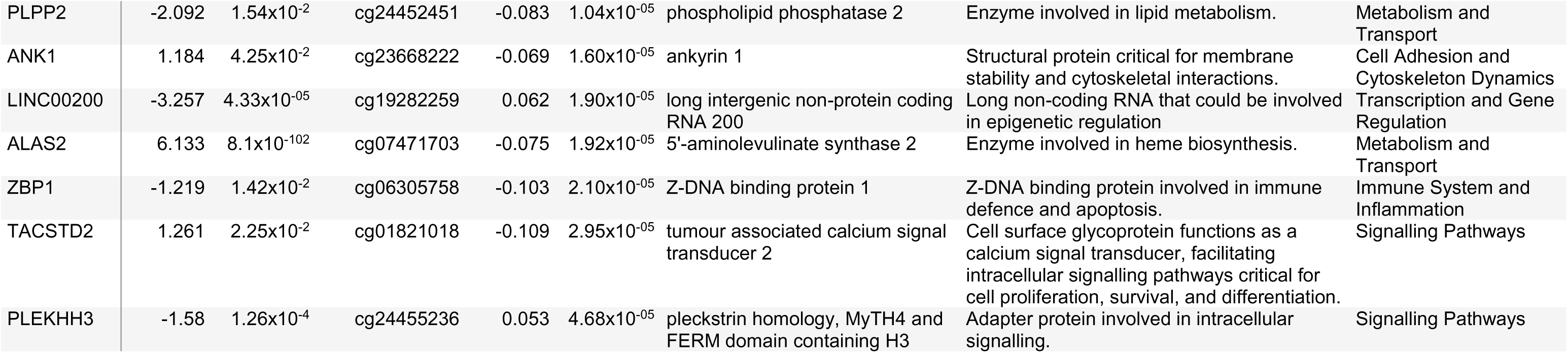
Significant methylome differences (p≤ 5 × 10^-5^) at baseline of sustained responders compared to non-responders together with their corresponding gene expression differences and associated molecular function.

| Gene Symbol | Expr fold change (rna) | Expr p-value (rna) | Illumina (cpG) | ΔBeta (cpG) | P-value (cpG) | Gene name | Description | Molecular Function |
| --- | --- | --- | --- | --- | --- | --- | --- | --- |
| DENND5B | -1.18 | $4.98 \times 10^{-2}$ | cg02046589 | 0.146 | $1.14 \times 10^{-34}$ | DENN domain containing 5B | Primarily involved in endosome dynamics, including the transport of vesicles. | Metabolism and Transport |
| ZFY | -1.407 | $9.48 \times 10^{-5}$ | cg00272582 | 0.15 | $1.14 \times 10^{-34}$ | zinc finger protein Y-linked | Zinc finger Y protein involved in transcriptional regulation and sex determination. | Transcription and Gene Regulation |
| PDGFRA | -1.405 | $4.56 \times 10^{-2}$ | cg21309167 | -0.203 | $3.59 \times 10^{-17}$ | platelet derived growth factor receptor alpha | Receptor tyrosine kinase involved in cell growth and development signalling. | Signalling Pathways |
| CPT1A | -1.468 | $1.37 \times 10^{-6}$ | cg10098373 | 0.139 | $3.68 \times 10^{-11}$ | carnitine palmitoyl transferase 1A | Carnitine palmitoyl transferase involved in fatty acid oxidation. | Metabolism and Transport |
| AHRR | -18.15 | $2.15 \times 10^{-19}$ | cg26076054 | -0.112 | $3.58 \times 10^{-10}$ | aryl hydrocarbon receptor repressor | Aryl hydrocarbon receptor repressor involved in immune response. | Immune System and Inflammation |
| RPH3AL | -1.222 | $3.41 \times 10^{-2}$ | cg17316718 | -0.12 | $1.07 \times 10^{-9}$ | rabphilin 3A like (without C2 domains) | Involved in synaptic vesicle recycling and signalling. | Neuronal/Brain Development and Function |
| CHI3L1 | 1.521 | $7.06 \times 10^{-7}$ | cg19081101 | -0.158 | $2.24 \times 10^{-9}$ | chitinase 3 like 1 | Chitinase-like protein involved in inflammation and tissue remodelling. | Immune System and Inflammation |
| UTY | -1.234 | $9.92 \times 10^{-3}$ | cg04790916 | 0.112 | $6.51 \times 10^{-7}$ | ubiquitously transcribed tetratricopeptide repeat, Y-linked | Y-chromosome gene involved in chromatin remodelling and gene expression | Epigenetics and Chromatin |
| LDHD | -1.373 | $1.39 \times 10^{-3}$ | cg00004883 | -0.041 | $8.35 \times 10^{-7}$ | lactate dehydrogenase D | Enzyme involved in lactate metabolism. | Metabolism and Transport |
| TBC1D16 | 1.204 | $3.73 \times 10^{-2}$ | cg26287152 | -0.095 | $1.4 \times 10^{-6}$ | TBC1 domain family member 16 | Regulates intracellular vesicle trafficking and signalling. | Signalling Pathways |
| FAM66A | 2.15 | $1.47 \times 10^{-2}$ | cg23285059 | -0.108 | $1.42 \times 10^{-6}$ | family sequence similarity 66 member A | Uncharacterised but implicated in viral infections. | Epigenetics and Chromatin |
| NME8 | 1.261 | $1.8 \times 10^{-2}$ | cg02531859 | 0.076 | $2.23 \times 10^{-6}$ | NME/NM23 family member 8 | NME8 is an essential structural and functional component of motile cilia and flagella. | Cell Adhesion and Cytoskeleton Dynamics |
| EIF1AY | -1.927 | $2.83 \times 10^{-14}$ | cg13308744 | 0.124 | $2.52 \times 10^{-6}$ | eukaryotic translation initiation factor 1A Y-linked | Translation initiation factor involved in protein synthesis regulation. | Transcription and Gene Regulation |
| PCBP3 | 1.323 | $1.06 \times 10^{-2}$ | cg13695288 | -0.114 | $4.01 \times 10^{-6}$ | poly(rC) binding protein 3 | RNA-binding protein involved in mRNA stability. | Transcription and Gene Regulation |
| PAQR6 | 1.397 | $4.36 \times 10^{-4}$ | cg03954786 | -0.124 | $6.09 \times 10^{-6}$ | progesterone adipoQ receptor family member 6 | Membrane receptor involved in signalling pathways. | Signalling Pathways |
| KCNK17 | -2.412 | $7.13 \times 10^{-10}$ | cg19475903 | 0.112 | $6.63 \times 10^{-6}$ | potassium two pore domain channel subfamily K member 17 | Potassium channel involved in membrane potential regulation. | Signalling Pathways |
| PLPP2 | -2.092 | 1.54x10<sup>-2</sup> | cg24452451 | -0.083 | 1.04x10<sup>-05</sup> | phospholipid phosphatase 2 | Enzyme involved in lipid metabolism. | Metabolism and Transport |
| ANK1 | 1.184 | 4.25x10<sup>-2</sup> | cg23668222 | -0.069 | 1.60x10<sup>-05</sup> | ankyrin 1 | Structural protein critical for membrane stability and cytoskeletal interactions. | Cell Adhesion and Cytoskeleton Dynamics |
| LINC00200 | -3.257 | 4.33x10<sup>-05</sup> | cg19282259 | 0.062 | 1.90x10<sup>-05</sup> | long intergenic non-protein coding RNA 200 | Long non-coding RNA that could be involved in epigenetic regulation | Transcription and Gene Regulation |
| ALAS2 | 6.133 | 8.1x10<sup>-102</sup> | cg07471703 | -0.075 | 1.92x10<sup>-05</sup> | 5'-aminolevulinate synthase 2 | Enzyme involved in heme biosynthesis. | Metabolism and Transport |
| ZBP1 | -1.219 | 1.42x10<sup>-2</sup> | cg06305758 | -0.103 | 2.10x10<sup>-05</sup> | Z-DNA binding protein 1 | Z-DNA binding protein involved in immune defence and apoptosis. | Immune System and Inflammation |
| TACSTD2 | 1.261 | 2.25x10<sup>-2</sup> | cg01821018 | -0.109 | 2.95x10<sup>-05</sup> | tumour associated calcium signal transducer 2 | Cell surface glycoprotein functions as a calcium signal transducer, facilitating intracellular signalling pathways critical for cell proliferation, survival, and differentiation. | Signalling Pathways |
| PLEKHH3 | -1.58 | 1.26x10<sup>-4</sup> | cg24455236 | 0.053 | 4.68x10<sup>-05</sup> | pleckstrin homology, MyTH4 and FERM domain containing H3 | Adapter protein involved in intracellular signalling. | Signalling Pathways |

Functional analysis utilising Gene Ontology of the 112 genes that were both differentially methylated and expressed at BAS between participants, who were found to either respond or not respond to ketamine therapy revealed the majority of influence was through the metabolic and transport pathways, alongside transcription and gene regulation pathways (**Table 4**).

**Table 4:**
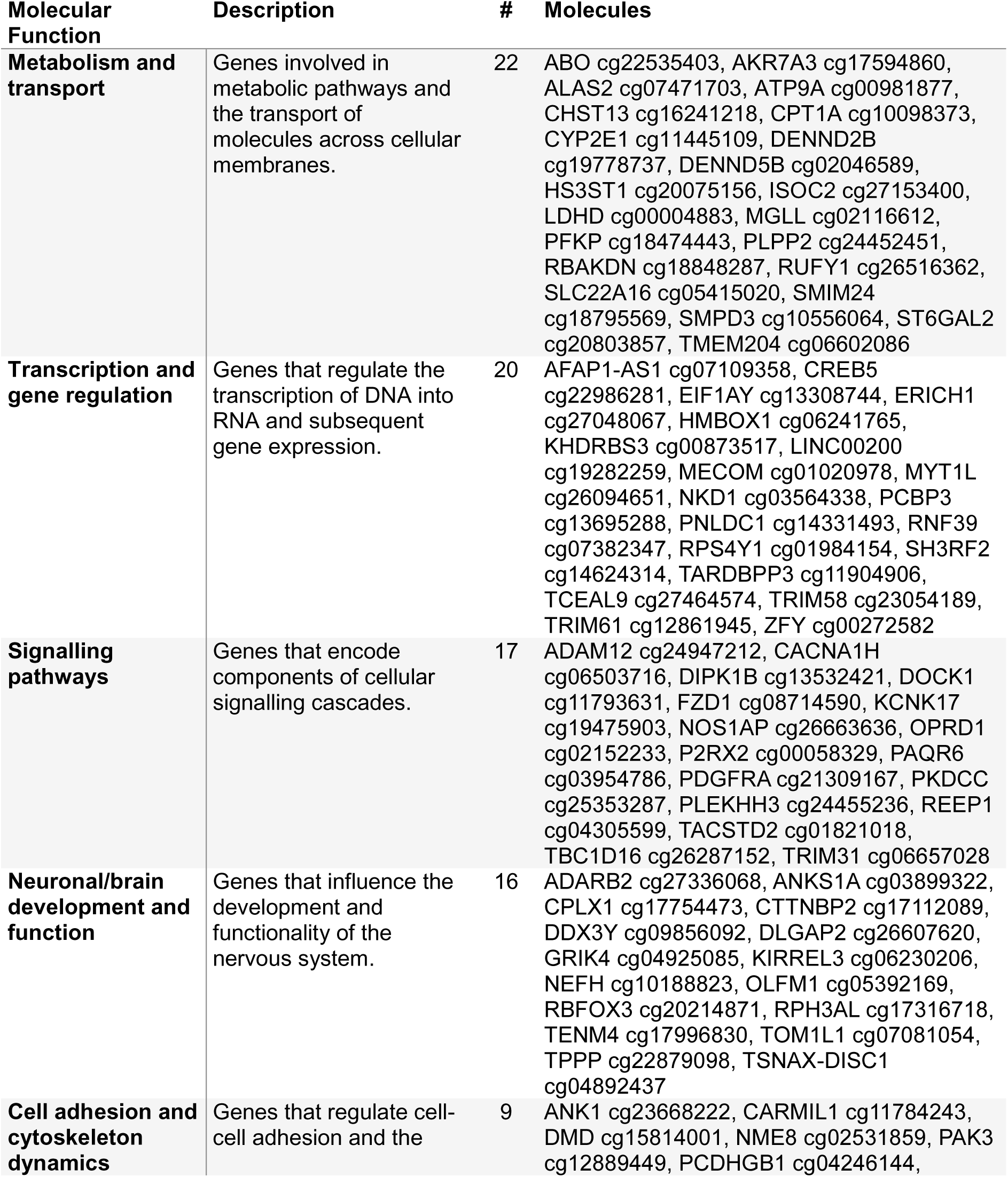
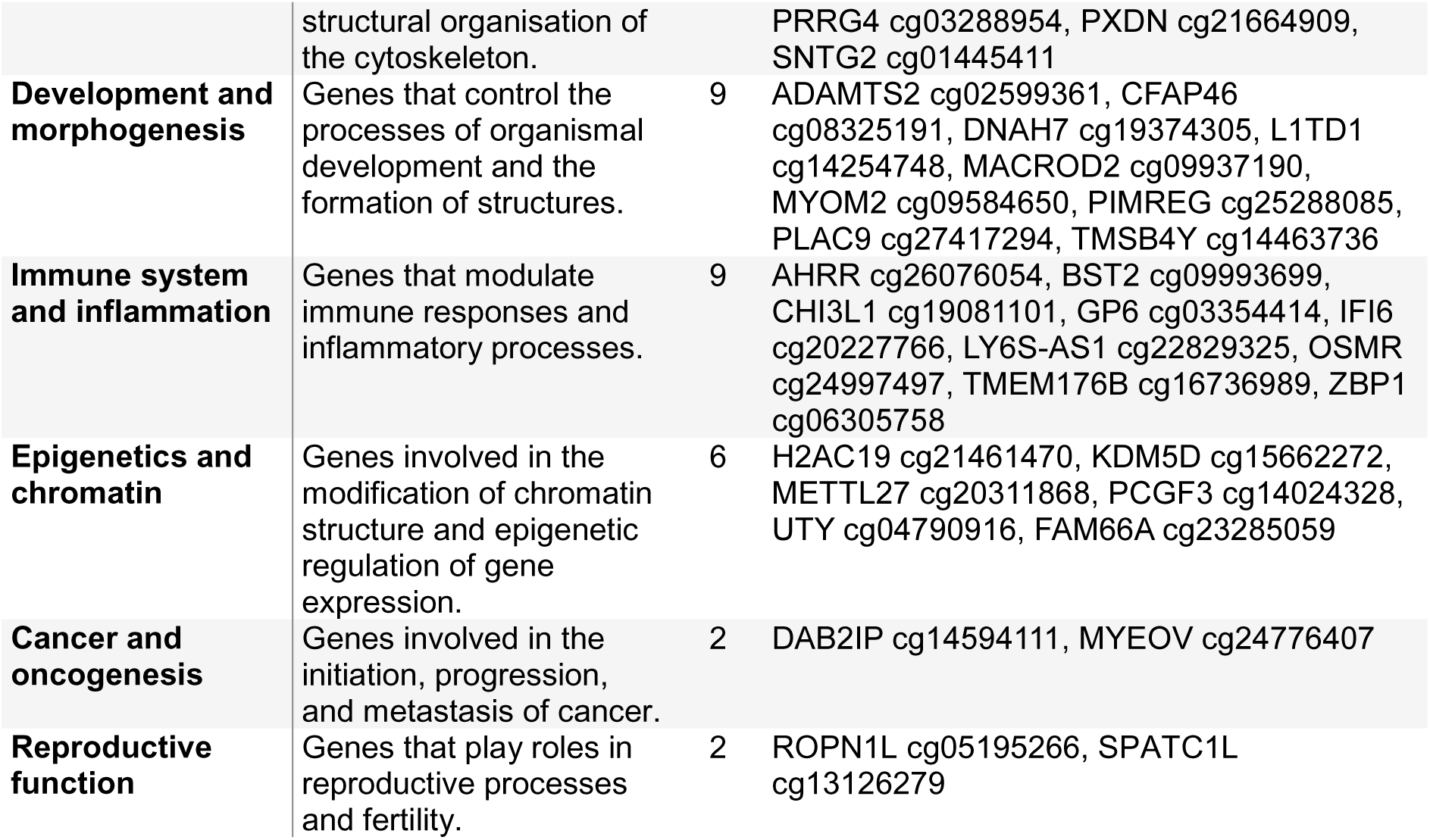
Overview of all 112 differentially methylated and expressed genes grouped according to molecular function.

| **Molecular Function** | **Description** | **#** | **Molecules** |
| --- | --- | --- | --- |
| **Metabolism and transport** | Genes involved in metabolic pathways and the transport of molecules across cellular membranes. | 22 | ABO cg22535403, AKR7A3 cg17594860, ALAS2 cg07471703, ATP9A cg00981877, CHST13 cg16241218, CPT1A cg10098373, CYP2E1 cg11445109, DENND2B cg19778737, DENND5B cg02046589, HS3ST1 cg20075156, ISOC2 cg27153400, LDHD cg00004883, MGLL cg02116612, PFKP cg18474443, PLPP2 cg24452451, RBAKDN cg18848287, RUFY1 cg26516362, SLC22A16 cg05415020, SMIM24 cg18795569, SMPD3 cg10556064, ST6GAL2 cg20803857, TMEM204 cg06602086 |
| **Transcription and gene regulation** | Genes that regulate the transcription of DNA into RNA and subsequent gene expression. | 20 | AFAP1-AS1 cg07109358, CREB5 cg22986281, EIF1AY cg13308744, ERICH1 cg27048067, HMBOX1 cg06241765, KHDRBS3 cg00873517, LINC00200 cg19282259, MECOM cg01020978, MYT1L cg26094651, NKD1 cg03564338, PCBP3 cg13695288, PNLDC1 cg14331493, RNF39 cg07382347, RPS4Y1 cg01984154, SH3RF2 cg14624314, TARDBPP3 cg11904906, TCEAL9 cg27464574, TRIM58 cg23054189, TRIM61 cg12861945, ZFY cg00272582 |
| **Signalling pathways** | Genes that encode components of cellular signalling cascades. | 17 | ADAM12 cg24947212, CACNA1H cg06503716, DIPK1B cg13532421, DOCK1 cg11793631, FZD1 cg08714590, KCNK17 cg19475903, NOS1AP cg26663636, OPRD1 cg02152233, P2RX2 cg00058329, PAQR6 cg03954786, PDGFRA cg21309167, PKDCC cg25353287, PLEKHH3 cg24455236, REEP1 cg04305599, TACSTD2 cg01821018, TBC1D16 cg26287152, TRIM31 cg06657028 |
| **Neuronal/brain development and function** | Genes that influence the development and functionality of the nervous system. | 16 | ADARB2 cg27336068, ANKS1A cg03899322, CPLX1 cg17754473, CTTNBP2 cg17112089, DDX3Y cg09856092, DLGAP2 cg26607620, GRIK4 cg04925085, KIRREL3 cg06230206, NEFH cg10188823, OLFM1 cg05392169, RBFOX3 cg20214871, RPH3AL cg17316718, TENM4 cg17996830, TOM1L1 cg07081054, TPPP cg22879098, TSNAX-DISC1 cg04892437 |
| **Cell adhesion and cytoskeleton dynamics** | Genes that regulate cell-cell adhesion and the | 9 | ANK1 cg23668222, CARMIL1 cg11784243, DMD cg15814001, NME8 cg02531859, PAK3 cg12889449, PCDHGB1 cg04246144, |
|  | structural organisation of the cytoskeleton. |  | PRRG4 cg03288954, PXDN cg21664909, SNTG2 cg01445411 |
| **Development and morphogenesis** | Genes that control the processes of organismal development and the formation of structures. | 9 | ADAMTS2 cg02599361, CFAP46 cg08325191, DNAH7 cg19374305, L1TD1 cg14254748, MACROD2 cg09937190, MYOM2 cg09584650, PIMREG cg25288085, PLAC9 cg27417294, TMSB4Y cg14463736 |
| **Immune system and inflammation** | Genes that modulate immune responses and inflammatory processes. | 9 | AHRR cg26076054, BST2 cg09993699, CHI3L1 cg19081101, GP6 cg03354414, IFI6 cg20227766, LY6S-AS1 cg22829325, OSMR cg24997497, TMEM176B cg16736989, ZBP1 cg06305758 |
| **Epigenetics and chromatin** | Genes involved in the modification of chromatin structure and epigenetic regulation of gene expression. | 6 | H2AC19 cg21461470, KDM5D cg15662272, METTL27 cg20311868, PCGF3 cg14024328, UTY cg04790916, FAM66A cg23285059 |
| **Cancer and oncogenesis** | Genes involved in the initiation, progression, and metastasis of cancer. | 2 | DAB2IP cg14594111, MYEOV cg24776407 |
| **Reproductive function** | Genes that play roles in reproductive processes and fertility. | 2 | ROPN1L cg05195266, SPATC1L cg13126279 |

## 3 Discussion

This is the first study to analyse the methylome and transcriptome profiles in combination from PTSD patients prior to undergoing a six-week low, dose ranging, sub-anaesthetic oral ketamine treatment. By combining the clinical outcomes of ketamine treatment at conclusion (∼4 weeks post-therapy), with pre-treatment molecular differences in PTSD participants, this study begins to uncover how epigenetic and transcriptional mechanisms may functionally contribute to the longer-term differential treatment outcomes among individuals with PTSD. This analysis also begins to identify key molecular markers that may aid in the prediction of treatment response. This raises a new and exciting possibility for a personalised medicine approach to the treatment of PTSD.

### 3.1 Clinical Variations within Participant Characteristics

The distinguishing factors within the BAS assessment found sustained responders initially presented with more severe PTSD scores (*t*(17) = -3.82, *p*= 0.001), but also less severe MDD results across cohorts (*t*(17) = 1.69, *p* = 0.11) and less overall well-being (*t*(17) = 3.22, *p* = 0.005) when compared to non-responders. This milder depression was further seen in the fact that only 63% of sustained responders presented with clinical MDD at all, compared to all the non-responders presenting with MDD (*p* = 0.02). This indicates that those who responded to the treatment were initially experiencing less depression, and lower well-being, than those who did not respond to ketamine therapy.

Sustained responders demonstrated a significant improvement, with an average reduction of 28 points in PCL-5 scores and 25 points in MADRS scores, bringing them below the clinical threshold for MDD. Additionally, these individuals showed a 36-point increase in well-being, as measured by the WHO-5 assessment, equating to a 1.5-fold improvement in overall well-being. These findings suggest that individuals responding to ketamine therapy experienced not only reduced PTSD symptoms but also improvements in co-morbid conditions such as MDD.

Although trauma type did not reach statistical significance, it showed a meaningful difference in response outcomes, with over 75% of non-responders reporting childhood trauma compared to only 36% of sustained responders. This suggests that the timing of trauma exposure across a participant’s lifespan, and possibly within developmentally sensitive periods, may play a role in the therapeutic outcome of ketamine, potentially influencing the magnitude of response within the neural pathways targeted by the treatment. Brain-derived neurotrophic factor (BDNF), a key neurotrophin, has been implicated in studies showing a connection between childhood trauma-induced hypomethylation and adult depression (12, 13). Another study also found epigenetic signatures in germ cells that are capable of responding to parental life stressors, thus shaping offspring neurodevelopment and potentially mediating transgenerational outcomes and altering stress reactivity in later life(31). These findings underscore the intricate relationship between baseline clinical presentation, mental health comorbidities, and trauma history in influencing the therapeutic response to ketamine, emphasising the importance of integrating clinical and molecular factors to develop precision therapies that optimise patient outcomes.

### 3.2 Biomarkers for the Sustained Response to Ketamine Treatment

As the cohort size was limited, combining methylome and transcriptome approaches enhanced the power of the analysis by increasing specificity, sensitivity, and functional relevance through high-dimensional data detection. This dual-layered strategy reduced false positives, deepened the understanding of molecular mechanisms, and linked biomarkers to biological pathways, thereby improving predictive power and pathway analysis. Of the 112 genes identified as statistically significant through this two-stage screening process, 48 genes exhibited unexpected methylation correlations, warranting further investigation in larger sample sizes to clarify their functional relevance and biomarker potential. The key differences observed in methylome and transcriptome data between sustained and non-responders revealed distinct molecular functions shaping predictive profiles of ketamine’s therapeutic efficacy. These differences are explored across several critical domains, including metabolism and transport, signalling pathways, transcriptional regulation, immune function, neuronal activity, epigenetics, and cytoskeletal dynamics, which underscores the multifaceted molecular landscape underpinning treatment response.

### 3.3 Metabolism and Transport

Metabolic and transport-related genes, including DENND5B cg02046589, LDHD cg00004883, and CPT1A cg10098373, emerged as strong candidates for predicting response to ketamine. DENND5B, which contributes to endosome activity(32), showed reduced expression and increased CpG methylation in sustained responders, possibly stabilising synaptic receptor activity and supporting neural plasticity, while non-responders exhibited ongoing disruptions in cellular signalling. Altered LDHD expression suggests a role in maintaining neuronal energy homeostasis under stress, with reduced activity disrupting lactate recycling, leading to energy deficits and impaired synaptic function(33). Similarly, the decreased expression of LDHD and CPT1A, enzymes central to lactate and fatty acid metabolism(34, 35), may have reflected a shift in sustained responders toward more efficient energy use, enhancing neural adaptability. Reduced CPT1A expression has been linked to decreased energy production, impairing neural activity and resilience under stress(36). Furthermore, metabolic deficits, including reduced fatty acid oxidation, are frequently observed in individuals with MDD, contributing to cognitive and emotional symptoms(37). Non-responders lacked this metabolic efficiency, potentially leading to energy-related neural dysfunction.

### 3.4 Signalling Pathways

Signalling pathways also emerged as significant factors in ketamine response. PLPP2 cg24452451, which regulates lipid metabolism, showed reduced expression in sustained responders, possibly facilitating neuronal membrane stabilisation. Non-responders, however, may have experienced adverse effects from elevated PLPP2 activity, which could disrupt lipid signalling and impede neural repair(38). Enhanced mitochondrial function in sustained responders is indirectly observed through the increased expression of ALAS2 cg07471703, a gene involved in heme biosynthesis(39). This improvement supports recovery from stress, a process that may be impaired in non-responders.

### 3.5 Transcription and Gene Regulation

Transcriptional regulation appears to play a role in the sustained effects of ketamine. The decreased expression of genes such as EIF1AY cg13308744, ZFY cg00272582, and PCBP3 cg13695288 in sustained responders may have stabilised RNA processing and reduced overactive protein synthesis, aligning with ketamine’s potential impact on cellular hyperactivity(40–42). In contrast, non-responders may be experienced prolonged transcriptional dysregulation, which may limit resilience and adaptation. Dysregulation of non-coding RNA, exemplified by a decreased expression in LINC00200 cg19282259 (C10orf139), may further compromise gene regulation in non-responders, while sustained responders benefited from adaptive epigenetic changes.

### 3.6 Immune System and Inflammation

Immune and inflammatory responses also differed. Dysregulated AHRR expression may prolong activation of stress-related immune pathways, exacerbating neuroinflammation, a hallmark of PTSD(43, 44). In sustained responders, substantial reductions in the expression of AHRR cg26076054 (associated with neuroinflammation) may have highlighted ketamine’s capacity to modulate immune pathways(43–45). Elevated CHI3L1 levels correlate with increased neuroinflammation in MDD and PTSD, acting as a potential compensatory mechanism to promote tissue repair following neural damage caused by chronic stress or depression (46). Increased CHI3L1 cg19081101 expression reflected anti-inflammatory properties and may be promoting tissue repair in sustained responders(47, 48). Non-responders, however, may have faced unresolved inflammation, seen with the dysregulated activity of ZBP1 cg06305758, which may exacerbate PTSD symptoms and impede treatment efficacy.

### 3.7 Neuronal/brain Development and Function

Genes linked to synaptic plasticity and signalling, including RPH3AL cg17316718, TBC1D16 cg26287152, and PDGFRA cg21309167, also show important differences (**Table 3**). The products of these genes regulate synaptic communication and repair(49–51). Altered PDGF signalling, a marker in postmortem brain tissue of individuals with MDD suggests a role in neuronal maintenance and stress response, with chronic stress models associating PDGFRA dysregulation with reduced neurogenesis and heightened inflammation(52). RPH3AL, involved in synaptic vesicle recycling, has been linked to decreased synaptic plasticity and impaired neurotransmitter release in animal models of depression, indicating its role in synaptic dysfunction in MDD (53). Sustained responders presented more favourable expression of these genes, which may lead to improved signalling and plasticity compared to non-responders. Enhanced neuro-steroid signalling through increased PAQR6 cg03954786 expression and the decreased expression of KCNK17 cg19475903 and TACSTD2 cg01821018 in sustained responders suggests that they may have benefited from stabilised neural activity and improved connectivity(54–56). Non-responders, however, exhibited continued disruptions in these pathways.

### 3.8 Epigenetics and Chromatin

UTY cg04790916 and FAM66A cg23285059 both had decreased expression and increased methylation in sustained responders indicating a higher degree of transcriptional stability may be present. UTY, a Y-chromosome gene involved a histone demethylase targeting H3K27me3(57), is known to regulate stress-responsive genes in males, and had increased methylation in sustained responders. Alternatively, higher expression levels of UTY in non-responders may have led to an inability to remodel chromatin adaptively, contributing to treatment resistance. Similarly, the decreased expression of FAM66A, involved in transcriptional regulation(58), suggested ketamine-induced epigenetic remodelling may promote transcriptional stabilisation, whereas non-responders may experience persistent dysregulation, impairing stress-related pathway modulation.

### 3.9 Cell Adhesion and Cytoskeleton Dynamics

NME8 cg02531859 associated with microtubule stabilisation and intracellular transport(59), had increased gene expression and reduced methylation in sustained responders, suggesting enhanced intracellular transport supporting neural plasticity may have been present. Alternatively, lower levels of NME8 in non-responders may have limited their ability to achieve structural and functional neuronal resilience. Similarly, ANK1 cg23668222 a structural protein essential for membrane stability and cytoskeletal organisation(60), has increased expression and reduced methylation in sustained responders, indicating improved cytoskeletal stability and synaptic connectivity. Differential methylation of ANK1 in individuals with MDD highlights its role in epigenetic regulation of structural and functional neuronal changes(61). Impaired ANK1 function is associated with reduced dendritic spine density and synaptic connectivity in depression(62). Conversely, the lowered levels of ANK1 in non-responders may have resulted in persistent membrane instability and impaired synaptic repair, thus reducing treatment efficacy.

These findings reveal significant differences in cellular states between sustained responders and non-responders, with ketamine influencing multiple domains, including metabolism, signalling pathways, gene regulation, immune response, neuronal function, and cytoskeletal dynamics. Sustained responders presented with metabolic differences linked to neural adaptability, improved signalling and plasticity, transcriptional stabilisation, and reduced inflammation. In contrast, non-responders exhibited persistent dysregulation across these pathways, which may hinder effective treatment outcomes. Notably, genes such as UTY and FAM66A highlighted epigenetic remodelling differences in responders, contributing to transcriptional stability, whereas non-responders faced ongoing disruptions. Additionally, the involvement of Y-linked genes in transcriptional regulation suggests potential sex-specific factors in treatment response.

### 3.10 Implications for Psychiatric Research

These findings highlight the need for longitudinal studies to track the dynamic nature of epigenetic and transcriptional responses throughout treatment, offering insights into recovery mechanisms and potential early markers of therapeutic response.

The integration of multi-omics approaches, including transcriptomics, methylomics, and proteomics, will be essential to fully elucidate the roles of these markers. Analysing the expression and methylation patterns within these biomarkers prior to treatment may provide insights into multi-target therapy to ensure the most effective treatment outcome across multiple conditions.

By combining detailed clinical presentation with comprehensive molecular profiling, there is significant potential for developing non-invasive diagnostic tools, such as blood or saliva-based biomarker panels, for monitoring treatment progress and predicting outcomes. Combining pharmacotherapeutics with ketamine treatment based on individual’s molecular profile, known functional impacts of targets and specific clinical presentation may also pave the way for more effective and sustained individualised management of psychiatric disorders.

Finally, personalised treatment strategies can leverage these markers to better tailor therapeutic interventions. For example, non-responders characterised by dysregulated PLPP2, and impaired lipid signalling may benefit from a combination therapy including phospholipase A2 (PLA2) inhibitors to offset potential inhibitory therapeutic effects(63). Those with altered PAQR6 expression might respond to therapy by combining two synthetic forms of allopregnanolone to enhance neuro-steroid signalling(64). Integrating these biomarkers with clinical characteristics and combining therapeutics could improve treatment precision and efficacy, reducing reliance on generalised therapeutic approaches.

## 4 Limitations and Future Directions

While this study has uncovered valuable insights, there are limitations to acknowledge. The small sample size limits the generalisability of the findings, and the lack of genetic diversity may limit the ability to extrapolate findings to non-Caucasian populations, a factor that future studies should address. Additionally, the observational nature of this study precludes causal inferences. Experimental approaches, such as gene knockdown or overexpression studies, may be necessary to establish the functional relevance of the identified biomarkers. Further, longitudinal studies tracking epigenetic and transcriptional changes over extended periods, whilst controlling for confounding factors, would also provide deeper insights into ketamine’s long-term therapeutic effects.

## 5 Conclusion

This study represents a significant step toward understanding the molecular underpinnings of ketamine’s sustained efficacy in PTSD treatment. By comparing the significant differences in sustained and non-responder characteristics before treatment, this study identified several key findings. Firstly, individuals who demonstrated a sustained response to ketamine treatment began treatment with higher PTSD symptom scores and lower MDD scores compared to non-responders. Secondly, the dose of ketamine required for a sustained response was on average lower than dosages given to non-responders, suggesting that a threshold may exist above which ketamine therapy may likely be unsuccessful. Finally, key methylation and gene expression biomarkers were identified that differentiated sustained responders from non-responders prior to treatment. These biomarkers are implicated in functionally relevant pathways which are positively correlated with treatment outcomes. These findings underscore the potential of integrating epigenetic and transcriptomic analyses to advance psychiatric research and improve clinical outcomes via a precision medicine approach for individuals with PTSD.

## Data Availability

All Illumina Human Methylation EPIC v2.0 Bead Chip raw and processed data has been registered with the Gene Expression Omnibus under accession number GSE287261, and Illumina RNA-Seq 20 million sequence paired end reads, raw and processed data alongside deidentified metadata has been registered with the Gene Expression Omnibus under GSE288174 and support the findings of this study. Deidentified behavioural scale results are available on request although original copies are protected for participant privacy.

## Acknowledgements

We would like to thank the Thompson Institute and the University of the Sunshine Coast for their support of molecular research into PTSD.

## Ethics approval and consent to participant

Participants were recruited from the Oral Ketamine Trial on Post-Traumatic Stress Disorder (OKTOP; Prince Charles Hospital HREC Approval: HREC/18/QPCH/288, UniSC Ethics Approval: A181190). Genetic analysis and reporting are registered under the Genetic Biomarkers of Ketamine on PTSD project (GBOK; Prince Charles Hospital HREC Approval: HREC/18/QPCH/288, UniSC Ethics Approval: S211655). Informed consent was obtained from patients over the age of 18 with written approval before commencement of the research.

## Declarations

This manuscript will contribute to a Doctorate by Research (PhD) from the University of the Sunshine Coast for author N.W.

## Author Contributions

Article conceptualisation, N.W., and A.K.; methodology, N.W. and A.K.; formal analysis, N.W. and A.K.; writing—original draft and editing, N.W.; writing—review and editing, A.K., supervision, A.K., A.B, B.Q, P.S and J.L. All authors have read and agreed to the published version of the manuscript.

## Author information

- National PTSD Research Centre, Thompson Institute, University of the Sunshine Coast (UniSC), Birtinya, QLD Australia Nathan J. Wellington, Bonnie L. Quigley, Ana P. Boucas, Paul E. Schwenn
- School of Health, UniSC, Sippy Downs, QLD Australia. Anna V. Kuballa, Nathan J. Wellington
- Centre for Bioinnovation, UniSC, Sippy Downs, QLD Australia Anna V. Kuballa, Bonnie L. Quigley, Nathan J. Wellington
- Sunshine Coast Health Institute, Sunshine Coast Hospital and Health Service, Birtinya, QLD Australia Bonnie L. Quigley, Nathan J. Wellington
- Thompson Brain and Mind Healthcare, Maroochydore, QLD Australia Jim Lagopoulos

## References

1. Tortora F, Hadipour AL, Battaglia S, Falzone A, Avenanti A, Vicario CM. The Role of Serotonin in Fear Learning and Memory: A Systematic Review of Human Studies. Brain Sci. 2023;13(8).

2. Cvrcek P. Side effects of ketamine in the long-term treatment of neuropathic pain. Pain Med. 2008;9(2):253–7.

3. Feder A, Costi S, Rutter SB, Collins AB, Govindarajulu U, Jha MK, et al. A Randomized Controlled Trial of Repeated Ketamine Administration for Chronic Posttraumatic Stress Disorder. Am J Psychiatry. 2021;178(2):193–202.

4. Krystal JH, Kavalali ET, Monteggia LM. Ketamine and rapid antidepressant action: new treatments and novel synaptic signaling mechanisms. Neuropsychopharmacology. 2024;49(1):41–50.

5. Sanacora G, Frye MA, McDonald W, Mathew SJ, Turner MS, Schatzberg AF, et al. A Consensus Statement on the Use of Ketamine in the Treatment of Mood Disorders. JAMA Psychiatry. 2017;74(4):399–405.

6. Feder A, Parides MK, Murrough JW, Perez AM, Morgan JE, Saxena S, et al. Efficacy of intravenous ketamine for treatment of chronic posttraumatic stress disorder: a randomized clinical trial. JAMA Psychiatry. 2014;71(6):681–8.

7. Du R, Han R, Niu K, Xu J, Zhao Z, Lu G, et al. The Multivariate Effect of Ketamine on PTSD: Systematic Review and Meta-Analysis. Front Psychiatry. 2022;13:813103.

8. Kawatake-Kuno A, Murai T, Uchida S. A Multiscale View of the Mechanisms Underlying Ketamine’s Antidepressant Effects: An Update on Neuronal Calcium Signaling. Front Behav Neurosci. 2021;15:749180.

9. Daskalakis NP, Rijal CM, King C, Huckins LM, Ressler KJ. Recent Genetics and Epigenetics Approaches to PTSD. Curr Psychiat Rep. 2018;20(5):30.

10. Turecki G, Meaney MJ. Effects of the Social Environment and Stress on Glucocorticoid Receptor Gene Methylation: A Systematic Review. Biol Psychiat. 2016;79(2):87–96.

11. Logue MW, Miller MW, Wolf EJ, Huber BR, Morrison FG, Zhou Z, et al. An epigenome-wide association study of posttraumatic stress disorder in US veterans implicates several new DNA methylation loci. Clin Epigenetics. 2020;12(1):46.

12. Bondar NP, Merkulova TI. Brain-derived neurotrophic factor and early-life stress: Multifaceted interplay. J Biosci. 2016;41(4):751–8.

13. Zhou A, Ancelin ML, Ritchie K, Ryan J. Childhood adverse events and BDNF promoter methylation in later-life. Front Psychiatry. 2023;14:1108485.

14. Krzyzewska IM, Ensink JBM, Nawijn L, Mul AN, Koch SB, Venema A, et al. Genetic variant in CACNA1C is associated with PTSD in traumatized police officers. Eur J Hum Genet. 2018;26(2):247–57.

15. Muhie S, Gautam A, Yang R, Misganaw B, Daigle BJ, Jr., Mellon SH, et al. Molecular signatures of post-traumatic stress disorder in war-zone-exposed veteran and active-duty soldiers. Cell Rep Med. 2023;4(5):101045.

16. Snijders C, Maihofer AX, Ratanatharathorn A, Baker DG, Boks MP, Geuze E, et al. Meta-analysis of longitudinal epigenome-wide association studies of military cohorts reveals multiple CpG sites associated with post-traumatic stress disorder. 2019.

17. Rutten BPF, Vermetten E, Vinkers CH, Ursini G, Daskalakis NP, Pishva E, et al. Longitudinal analyses of the DNA methylome in deployed military servicemen identify susceptibility loci for post-traumatic stress disorder. Mol Psychiatry. 2018;23(5):1145–56.

18. Hawn SE, Neale Z, Wolf EJ, Zhao X, Pierce M, Fein-Schaffer D, et al. Methylation of the AIM2 gene: An epigenetic mediator of PTSD-related inflammation and neuropathology plasma biomarkers. Depress Anxiety. 2022;39(4):323–33.

19. Smith AK, Conneely KN, Kilaru V, Mercer KB, Weiss TE, Bradley B, et al. Differential immune system DNA methylation and cytokine regulation in post-traumatic stress disorder. Am J Med Genet B Neuropsychiatr Genet. 2011;156B(6):700–8.

20. Nothling J, Abrahams N, Toikumo S, Suderman M, Mhlongo S, Lombard C, et al. Genome-wide differentially methylated genes associated with posttraumatic stress disorder and longitudinal change in methylation in rape survivors. Transl Psychiat. 2021;11(1):594.

21. Dean KR, Hammamieh R, Mellon SH, Abu-Amara D, Flory JD, Guffanti G, et al. Multi-omic biomarker identification and validation for diagnosing warzone-related post-traumatic stress disorder. Mol Psychiatry. 2020;25(12):3337–49.

22. Siegel CE, Laska EM, Lin Z, Xu M, Abu-Amara D, Jeffers MK, et al. Utilization of machine learning for identifying symptom severity military-related PTSD subtypes and their biological correlates. Transl Psychiat. 2021;11(1):227.

23. Quigley BL, Can AT, Dutton M, Gallay CC, Forsyth G, Jones M, et al. Low dose oral ketamine treatment on post-traumatic stress disorder (PTSD) (OKTOP): An open-label pilot study. medRxiv. 2024:2024.11.26.24318024.

24. Weathers FW, Blake, D.D., Schnurr, P.P., Kaloupek, D.G., Marx, B.P., & Keane, T.M. . The Clinician-Administered PTSD Scale for DSM-5 (CAPS-5). 2013.

25. Villanueva RAM, Chen ZJ. ggplot2: Elegant Graphics for Data Analysis (2nd ed.). Measurement: Interdisciplinary Research and Perspectives. 2019;17(3):160–7.

26. Team R. RStudio: Integrated Development for R. Boston, MA: RStudio, PBC; 2020.

27. Ritchie ME, Phipson B, Wu D, Hu Y, Law CW, Shi W, et al. limma powers differential expression analyses for RNA-sequencing and microarray studies. Nucleic acids research. 2015;43(7):e47-e.

28. Vaughan HWaRFaLHaKMaD. dplyr: A Grammar of Data Manipulation. 2023.

29. Aryee MJ, Jaffe AE, Corrada-Bravo H, Ladd-Acosta C, Feinberg AP, Hansen KD, et al. Minfi: a flexible and comprehensive Bioconductor package for the analysis of Infinium DNA methylation microarrays. Bioinformatics. 2014;30(10):1363–9.

30. Thomas PD, Hill DP, Mi H, Osumi-Sutherland D, Van Auken K, Carbon S, et al. Gene Ontology Causal Activity Modeling (GO-CAM) moves beyond GO annotations to structured descriptions of biological functions and systems. Nat Genet. 2019;51(10):1429–33.

31. Rodgers AB, Morgan CP, Leu NA, Bale TL. Transgenerational epigenetic programming via sperm microRNA recapitulates effects of paternal stress. Proc Natl Acad Sci U S A. 2015;112(44):13699–704.

32. Gordon SM, Neufeld EB, Yang Z, Pryor M, Freeman LA, Fan X, et al. DENND5B Regulates Intestinal Triglyceride Absorption and Body Mass. Sci Rep-Uk. 2019;9(1):3597.

33. Herbet M, Natorska-Chomicka D, Korga A, Ostrowska M, Izdebska M, Gawronska-Grzywacz M, et al. Altered expression of genes involved in brain energy metabolism as adaptive responses in rats exposed to chronic variable stress; changes in cortical level of glucogenic and neuroactive amino acids. Mol Med Rep. 2019;19(3):2386–96.

34. Kwong AK, Wong SS, Rodenburg RJT, Smeitink J, Chan GCF, Fung CW. Human d-lactate dehydrogenase deficiency by LDHD mutation in a patient with neurological manifestations and mitochondrial complex IV deficiency. JIMD Rep. 2021;60(1):15–22.

35. Schlaepfer IR, Joshi M. CPT1A-mediated Fat Oxidation, Mechanisms, and Therapeutic Potential. Endocrinology. 2020;161(2):bqz046.

36. Schlaepfer IR, Joshi M. CPT1A-mediated Fat Oxidation, Mechanisms, and Therapeutic Potential. Endocrinology. 2020;161(2).

37. Chen H, Wang J, Chen S, Chen X, Liu J, Tang H, et al. Abnormal energy metabolism, oxidative stress, and polyunsaturated fatty acid metabolism in depressed adolescents associated with childhood maltreatment: A targeted metabolite analysis. Psychiatry Res. 2024;335:115795.

38. Zeng Y, Cao S, Li N, Tang J, Lin G. Identification of key lipid metabolism-related genes in Alzheimer’s disease. Lipids Health Dis. 2023;22(1):155.

39. Harigae H, Nakajima O, Suwabe N, Yokoyama H, Furuyama K, Sasaki T, et al. Aberrant iron accumulation and oxidized status of erythroid-specific delta-aminolevulinate synthase (ALAS2)-deficient definitive erythroblasts. Blood. 2003;101(3):1188–93.

40. Godfrey AK, Naqvi S, Chmatal L, Chick JM, Mitchell RN, Gygi SP, et al. Quantitative analysis of Y-Chromosome gene expression across 36 human tissues. Genome Res. 2020;30(6):860–73.

41. Koopman P, Ashworth A, Lovell-Badge R. The ZFY gene family in humans and mice. Trends Genet. 1991;7(4):132–6.

42. Pesic V, Petrovic J, M MJ. Molecular Mechanism and Clinical Relevance of Ketamine as Rapid-Acting Antidepressant. Drug Dev Res. 2016;77(7):414–22.

43. Smith AK, Ratanatharathorn A, Maihofer AX, Naviaux RK, Aiello AE, Amstadter AB, et al. Epigenome-wide meta-analysis of PTSD across 10 military and civilian cohorts identifies methylation changes in AHRR. Nat Commun. 2020;11(1):5965.

44. Nomura S, Morita H. Dysregulation of DNA Methylation in the Aryl-Hydrocarbon Receptor Repressor (AHRR) Gene. Circ J. 2022;86(6):993–4.

45. Garrett ME, Dennis MF, Bourassa KJ, Workgroup VAM-AM, Hauser MA, Kimbrel NA, et al. Genome-wide DNA methylation analysis of cannabis use disorder in a veteran cohort enriched for posttraumatic stress disorder. Psychiatry Res. 2024;333:115757.

46. Jiang W, Zhu F, Xu H, Xu L, Li H, Yang X, et al. CHI3L1 signaling impairs hippocampal neurogenesis and cognitive function in autoimmune-mediated neuroinflammation. Science Advances. 2023;9(39):eadg8148.

47. Jiang W, Zhu F, Xu H, Xu L, Li H, Yang X, et al. CHI3L1 signaling impairs hippocampal neurogenesis and cognitive function in autoimmune-mediated neuroinflammation. Sci Adv. 2023;9(39):eadg8148.

48. Mizoguchi E, Sadanaga T, Nanni L, Wang S, Mizoguchi A. Recently Updated Role of Chitinase 3-like 1 on Various Cell Types as a Major Influencer of Chronic Inflammation. Cells. 2024;13(8):678.

49. Hodgkinson CA, Enoch MA, Srivastava V, Cummins-Oman JS, Ferrier C, Iarikova P, et al. Genome-wide association identifies candidate genes that influence the human electroencephalogram. Proc Natl Acad Sci U S A. 2010;107(19):8695–700.

50. Bassil K, Ali N, Pishva E, van den Hove DL. Epigenome-Wide Association Studies in Psychiatry: Achievements and Problems. Epigenetic Epidemiology: Springer; 2022. p. 427–44.

51. Lewandowski SA, Fredriksson L, Lawrence DA, Eriksson U. Pharmacological targeting of the PDGF-CC signaling pathway for blood-brain barrier restoration in neurological disorders. Pharmacol Ther. 2016;167:108–19.

52. Alnafisah R, Alganem K, Hamoud A-r, Imami AS, Creeden J, Rayn V W, et al. P307. Dysregulated Kinase Networks in Major Depressive Disorder. Biological Psychiatry. 2022;91(9):S211–S2.

53. Aleksandrova LR, Wang YT, Phillips AG. Evaluation of the Wistar-Kyoto rat model of depression and the role of synaptic plasticity in depression and antidepressant response. Neurosci Biobehav Rev. 2019;105:1–23.

54. Pang Y, Dong J, Thomas P. Characterization, neurosteroid binding and brain distribution of human membrane progesterone receptors δ and ɛ (mPRδ and mPRɛ) and mPRδ involvement in neurosteroid inhibition of apoptosis. Endocrinology. 2013;154(1):283–95.

55. Lee LM, Muntefering T, Budde T, Meuth SG, Ruck T. Pathophysiological Role of K(2P) Channels in Human Diseases. Cell Physiol Biochem. 2021;55(S3):65–86.

56. Gouwens NW, Sorensen SA, Baftizadeh F, Budzillo A, Lee BR, Jarsky T, et al. Integrated Morphoelectric and Transcriptomic Classification of Cortical GABAergic Cells. Cell. 2020;183(4):935–53 e19.

57. Shpargel KB, Sengoku T, Yokoyama S, Magnuson T. UTX and UTY demonstrate histone demethylase-independent function in mouse embryonic development. PLoS Genet. 2012;8(9):e1002964.

58. Wong A, Zhou A, Cao X, Mahaganapathy V, Azaro M, Gwin C, et al. MicroRNA and MicroRNA-Target Variants Associated with Autism Spectrum Disorder and Related Disorders. Genes (Basel). 2022;13(8):1329.

59. Liu Y, Yu JT, Wang HF, Hao XK, Yang YF, Jiang T, et al. Association between NME8 locus polymorphism and cognitive decline, cerebrospinal fluid and neuroimaging biomarkers in Alzheimer’s disease. PLoS One. 2014;9(12):e114777.

60. Courtnell K. Immunohistochemical analysis of Ank1 in Tg4510 and J20 mouse models: University of Exeter (United Kingdom); 2022.

61. Zheng J, Womer FY, Tang L, Guo H, Zhang X, Tang Y, et al. Integrative omics analysis reveals epigenomic and transcriptomic signatures underlying brain structural deficits in major depressive disorder. Transl Psychiat. 2024;14(1):17.

62. Tang L, Zhao P, Pan C, Song Y, Zheng J, Zhu R, et al. Epigenetic molecular underpinnings of brain structural-functional connectivity decoupling in patients with major depressive disorder. J Affect Disord. 2024;363:249–57.

63. Chang J, Musser JH, McGregor H. Phospholipase A2: function and pharmacological regulation. Biochem Pharmacol. 1987;36(15):2429–36.

64. Genazzani AR, Petraglia F, Bernardi F, Casarosa E, Salvestroni C, Tonetti A, et al. Circulating levels of allopregnanolone in humans: gender, age, and endocrine influences. J Clin Endocrinol Metab. 1998;83(6):2099–103.

